# Application of machine learning and complex network measures to an EEG dataset from ayahuasca experiments

**DOI:** 10.1101/2022.05.31.22275817

**Authors:** Caroline L. Alves, Rubens Gisbert Cury, Kirstin Roster, Aruane M. Pineda, Francisco A. Rodrigues, Christiane Thielemann, Manuel Ciba

**Affiliations:** University of São Paulo (USP), Institute of Mathematical and Computer Sciences (ICMC), São Paulo, Brazil; BioMEMS lab, Aschaffenburg University of Applied Sciences, Aschaffenburg, Germany; University of São Paulo, Movement Disorders Center, Department of Neurology

## Abstract

Ayahuasca is made from a mixture of Amazonian herbs and has been used for a few hundred years by the people of this region for traditional medicine. In addition, this plant has been shown to be a potential treatment for various neurological and psychiatric disorders.

EEG experiments have found specific brain regions that changed significantly due to ayahuasca. Here, we used an EEG dataset to investigate the ability to automatically detect changes in brain activity using machine learning and complex networks. Machine learning was applied at three different levels of data abstraction: (A) the raw EEG time series, (B) the correlation of the EEG time series, and (C) the complex network measures calculated from (B).

As a result, the machine learning method was able to automatically detect changes in brain activity, with case (B) showing the highest accuracy (92%), followed by (A) (88%) and (C) (83%), indicating that connectivity changes between brain regions are more important than connectivity changes within brain regions. The most activated areas were the frontal and temporal lobe, which is consistent with the literature.

In terms of brain connections, the correlation between F3 and PO4 was the most important. This connection may point to a cognitive process similar to face recognition in individuals during ayahuasca-mediated visual hallucinations.

Furthermore, closeness centrality and assortativity were the most important complex network measures. These two measures are also associated with diseases such as Alzheimer’s disease, indicating a possible therapeutic mechanism.

Overall, our results showed that machine learning methods were able to automatically detect changes in brain activity during ayahuasca consumption. The results also suggest that the application of machine learning and complex network measurements are useful methods to study the effects of ayahuasca on brain activity and medical use.

## 1. INTRODUCTION

Ayahuasca is made from a blend of Amazonian herbs [1]. This combination of plants is often associated with rituals of different religions and social groups. Ayahuasca has been used in the Amazon for a couple of hundred years, being part of the traditional medicine of the natives within this region [2].

Since the use of ayahuasca has spread throughout many countries, it is necessary to study in depth its cerebral mechanisms and its potential clinical implications. In addition, because it affects brain areas related to emotions, memories, and executive functions, ayahuasca might be used in the treatment of psychiatric disorders, such as drug addiction [3–5], Parkinson’s disease [6–9], and depression [10] [11–16]. For example, an open-label clinical study found significant therapeutic benefits among patients with treatment-resistant major depressive disorder after the administration of a single dose of ayahuasca [12]. Moreover, a randomized trial showed that ayahuasca doses were associated with reductions in depressive symptoms in patients with major depressive disorder, compared to placebo treatments [11]. Additionally, ayahuasca has been shown to elicit antineuroinflammatory properties [16] and stimulate adult neurogenesis in vitro [17]. In this line, ayahuasca could be helpful for the treatment of several neurological diseases well known to harbor inflammation in its physiopathology [18], including chronic degenerative diseases and illnesses related to acute injury, such as cerebral ischemia, multiple sclerosis, and Alzheimer’s disease (AD) [19, 20]. However, in most countries ayahuasca is an illegal substance and only legalized for religious use only, like e.g. Brazil.

The application of mathematical methods of graph theory yielded interesting insights into the complex network structure of the human brain, which is also associated with pathological states [21–24]. Notably, complex networks parameters have been used as biomarker for several diseases [25, 26]. In this context, machine learning has been used for more accurate and automatic medical diagnosis [27–37] and may be an important tool capable of detecting acute and permanent abnormalities in the brain.

The present work aims to investigate whether complex network measurements combined with machine learning algorithms are suitable to detect temporal changes in the brain functionality of participants after consumption of ayahuasca. Specifically, using both exploratory and predictive approaches, this study raised the following research questions:

- Is it possible to automatically detect the changes in the brain activity after intake of ayahuasca with machine learning methods using the following data abstraction levels for the input: (A) raw EEG time series, (B) the correlation between the EEG electrodes as used in (A) represented by a connectivity matrix, and (C) complex network measures extracted from (B).
- Which data abstraction level is most important to detect the changes?
- What are important features that may serve as biomarker to detect the changes?

## II. DATA

The data ^1^ used for this study has been made open available by the Federal University of S ∼ao Carlos, Brazil [39]. Sixteen healthy male and female patients volunteered to drink ayahuasca while EEG recordings were performed. Patients were asked to remain in a resting-state with eyes closed. A nurse accompanied the experiment for its duration of 225 minutes. Recordings started 25 minutes before ingestion of ayahuasca and continued 200 minutes after ingestion. The main compounds in the brew were [39]: Dimethyltryptamine (DMT), DMT-N-oxide (DMT-NO), N-methyltryptamine (NMT), indoleacetic acid (IAA), 5-hydroxy-DMT (5-OH-DMT, or bufotenin), 5-methoxy-DMT (5-MeO-DMT), Harmine, Harmol, Harmaline, Harmalol, THH, 7-hydroxy-tetrahydroharmine (THH-OH), and 2-methyl-tetrahydro-beta- carboline (2-MTHBC). All recordings were downsampled to 500 Hz, bandpass filtered between 0.5 and 150 Hz and bad segments due to movements were removed. Recordings were made with 62 electrodes, following the EEG electrode positions in the 10-10 system. These channels are: Fp1, Fz, F3, F7, FT9, FC5, FC1, C3, TP9, CP5, CP1, Pz, P3, P7, O1, Oz, P8, TP10, CP6, CP2, C4, T8, FT10, FC6, FC2, F4, F8, Fp2, AF7, AF3, AFz, F1, F5, FT7, FC3, FCz, C1, C5, TP7, CP3, P1, P5, PO7, PO3, POz, PO4, PO8, P6, P2, CPz, CP4, TP8, FC4, FT8, F6, F2, AF4, AF8, O2, P4, C6, and C2 (see in appendix Figure A). Further details are given in [39].

## III. INPUT DATA FOR MACHINE LEARNING

The following three data abstraction levels were applied to a classification algorithm as described in subsection IV: (III A) the EEG time series (Figure 1), (III B) the connectivity matrix calculated by means of the Pearson correlation of the EEG time series (Figure 3), and (III C) the complex network measures calculated from the connectivity matrix (Figure 2).

**FIG. 1:**
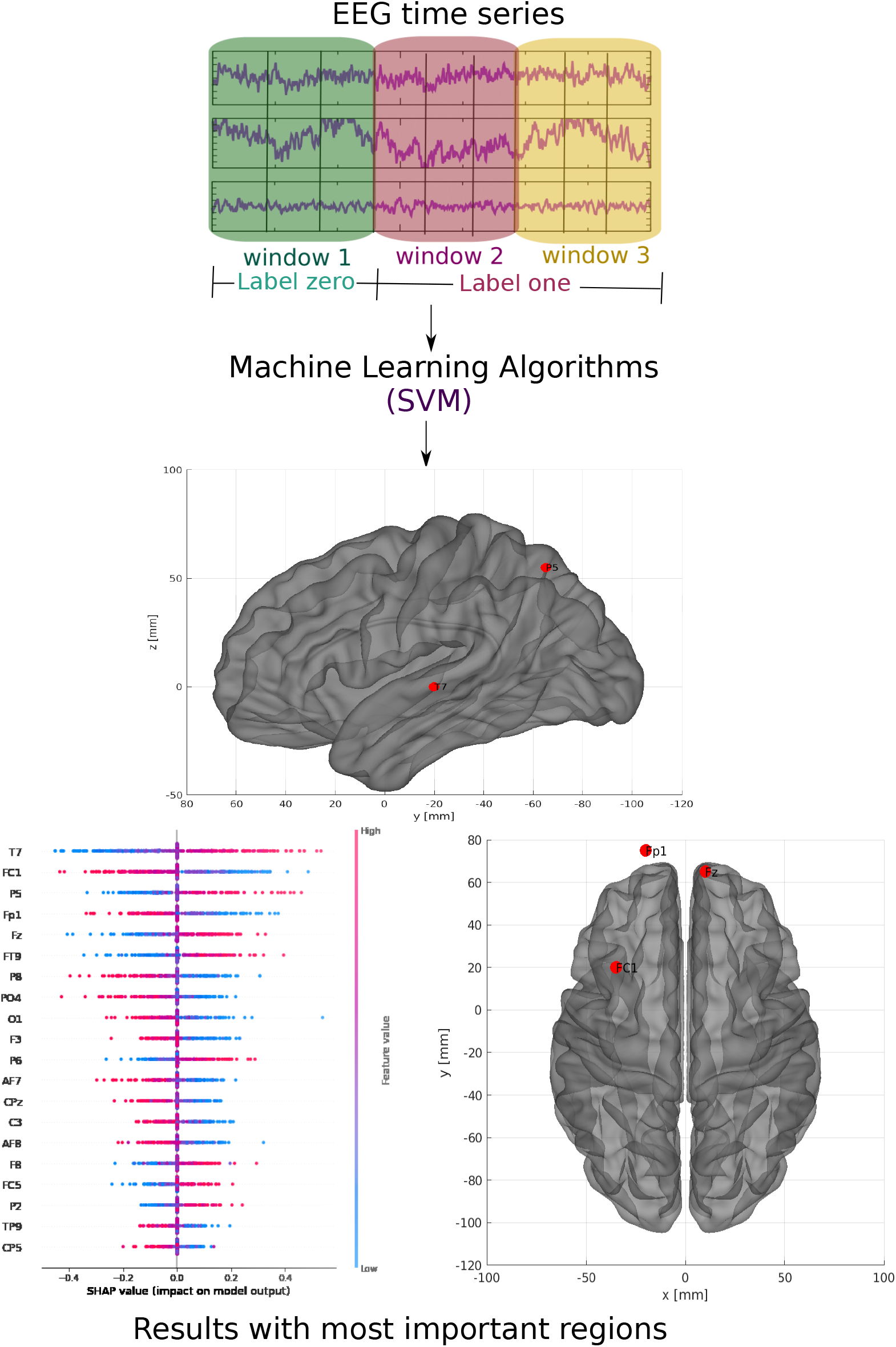
Methodology of the subsection III A using raw EEG time series. The EEG time series was split into three parts for all subjects. Those corresponding to the first window were labeled as 0 (no effect of ayahuasca) and those corresponding to the second and third windows as 1 (under the influence of ayahuasca) and then SVM was used.

**FIG. 2:**
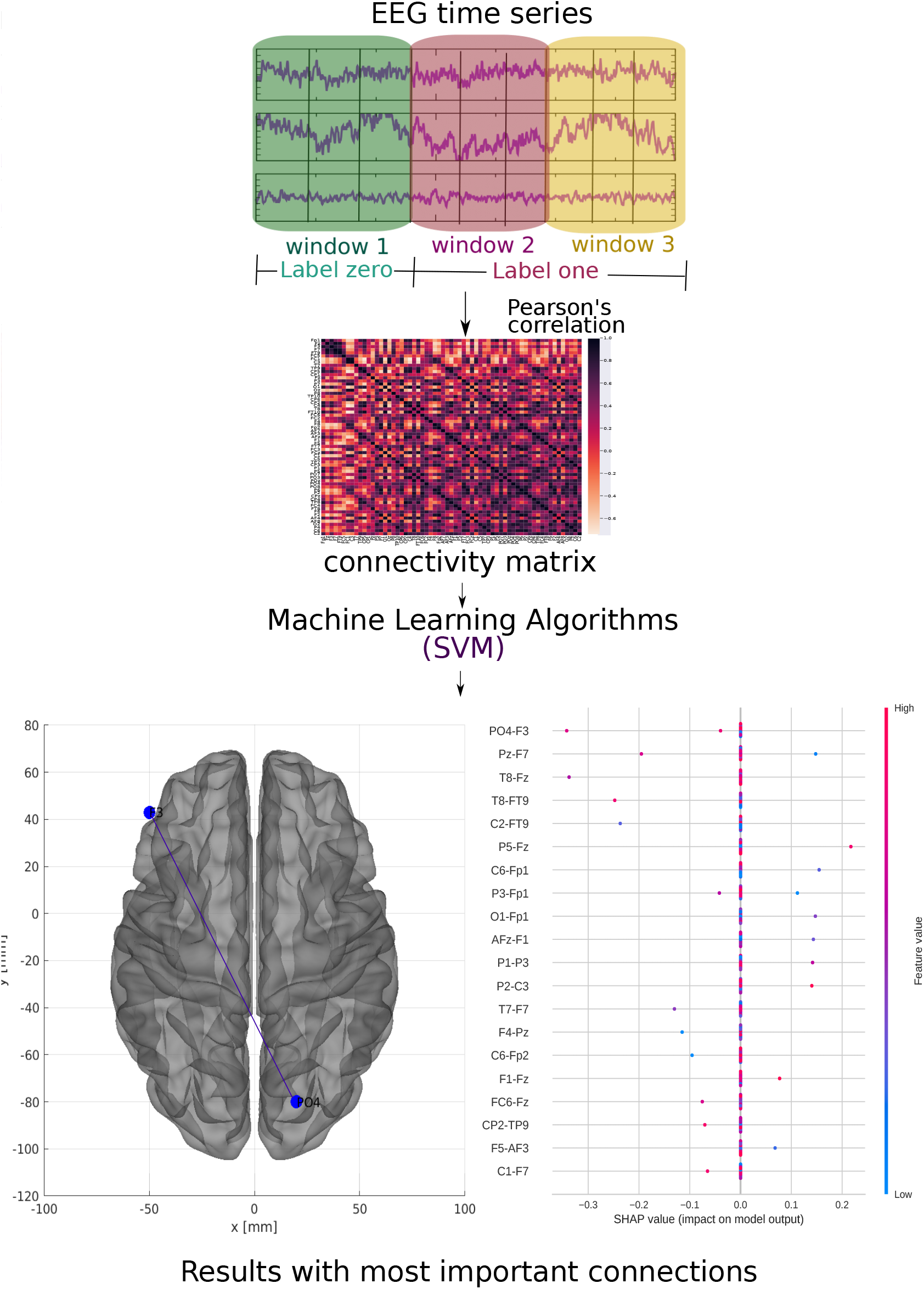
Methodology of the subsection III B using connectivity matrices. For each of the time windows, the Pearson correlation connectivity matrix was generated and then they were classified with the SVM method considering the first window as zero label (without ayahuasca) and the other two as one label (with ayahuasca). This analysis aimed to verify the best connections of the brain areas used during ayahuasca use.

**FIG. 3:**
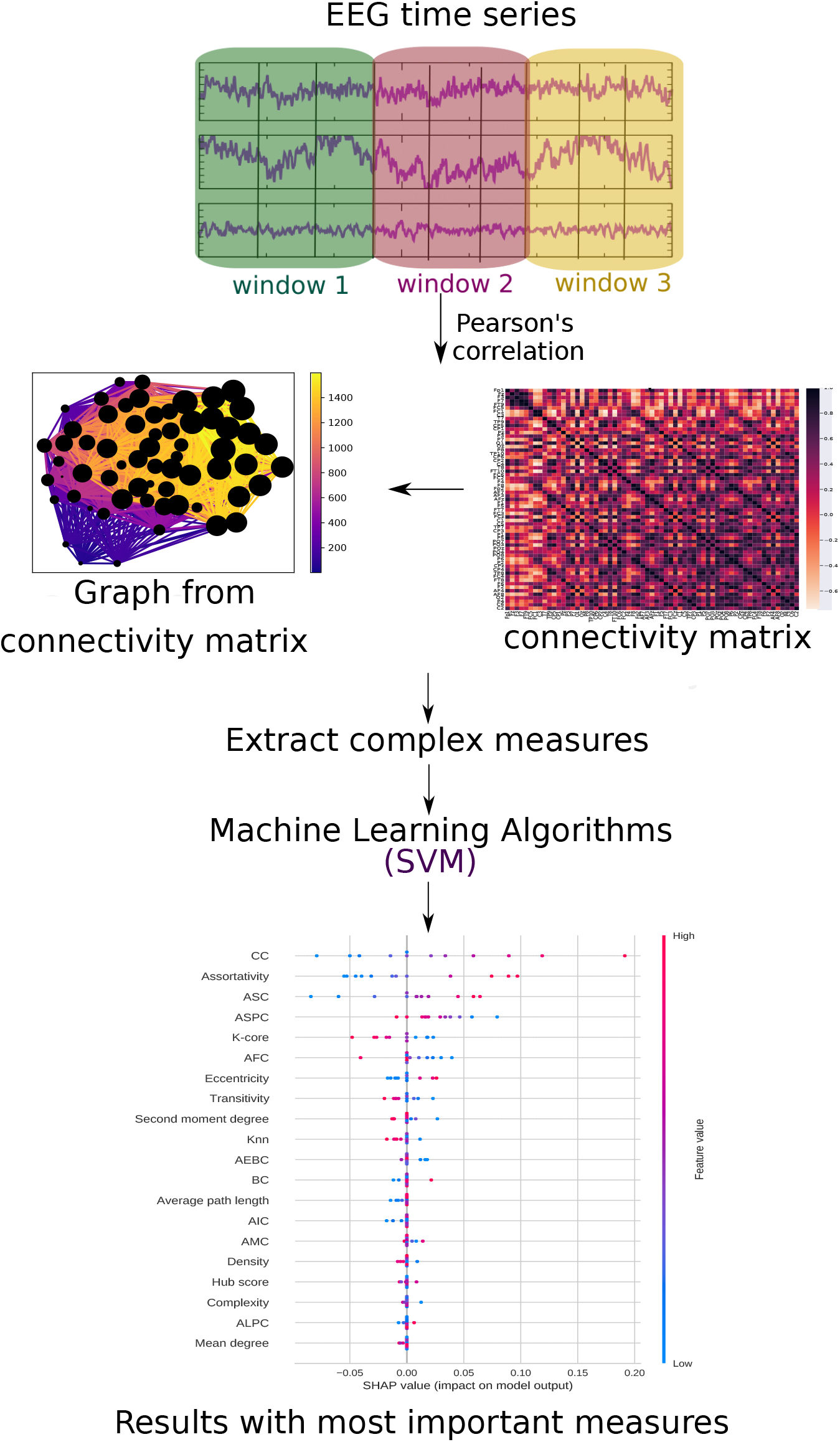
Methodology of the subsection III C using complex network measures. The EEG time series is divided into three parts. For each of them, Pearson correlation was calculated. For each window, a connectivity matrix was generated (in the Figure, the connectivity matrix of the first window of the first subject containing the 62 electrodes, the color bar contains the connection strength between these electrodes). For each of them, a graph was constructed (in the Figure, the graph of this connectivity matrix contains 62 nodes and the connection strength according to the color bar and the size of the node according to its number of connections) and from them complex measures of the network are extracted. These measurements were then evaluated with machine learning algorithm to see which one captures the changes in the brain with ayahuasca use.

### A. EEG time series

The data was divided into three “time windows” (see Table I). The first window (25 minutes before ingestion until 50 minutes after ingestion of ayahuasca) was defined as the “control”. This is reasonable as it is known from Schenberg *et al*. [39], that the blood plasma concentration of the main psychedelic compound DMT is low until 50 minutes after ingestion. Windows two and three were both defined as fully influenced by ayahuasca. The reason for splitting the ayahuasca-influenced time series into two windows was to increase the number of data points for the machine learning algorithm. Even though the number of independent samples (subjects) did not change, increasing the data points by splitting the time series is a common machine learning approach [40, 41]. In the following classification task, there will be only two classes with label zero (without ayahuasca) and label one (with ayahuasca). The scheme of this methodology is shown in Figure 1. To input the data into the machine learning algorithm, the EEG time series of all subjects were merged sequentially and stored in a 2D matrix. Each column represents an electrode and each row represents the amplitude of each time point of the EEG signal. Such a 2D matrix was generated for each of the three time windows.

**TABLE I:** Definition of time windows of the EEG signal. Window 1 is considered the control (without effect of ayahuasca), window 2 and 3 are considered as recordings under the influence of ayahuasca.

| Time window | Ingestion of ayahuasca<br>at $t = 0$ minutes |
| --- | --- |
| 1 | -25 to 50 minutes |
| 2 | 50 to 125 minutes |
| 3 | 125 to 200 minutes |

### B. Connectivity matrices

The matrices of connectivity were calculated by the well known Pearson correlation. It is a widely used and successfully approved measure to capture the correlation The goal was to identify the brain regions most affected by ayahuasca use. of EEG electrodes [42–46]. The Pearson correlation between was calculated for all electrode pairs resulting in three connectivity matrices per participant (for each time window). Figure 2 illustrates the workflow of this approach. To input the data into the machine learning algorithm, the connectivity matrices were flattened into one vector. Then, all vectors were sequentially merged into a 2D matrix. Each column represents a connection between two brain regions and each row represents a subject. Such a 2D matrix was generated for each of the three time windows.

### C. Complex network measures

For each connectivity matrix (see subsection III B), a graph was generated to extract different complex network measures. To input the data into the machine learning algorithm, the complex network measures were stored in a matrix. Each column represents a complex network measure and each row a subject. Such a 2D matrix was generated for each of the three time windows. The following complex network measures were calculated: Assortativity [47, 48], average path length [49], betweenness centrality (BC) [50], closeness centrality (CC) [51], eigenvector centrality (EC) [52], diameter [53], hub score [54], average degree of nearest neighbors [55] (Knn), mean degree [56], second moment degree [57], entropy degree [58], transitivity [59, 60], complexity, k-core [61, 62], eccentricity [63], density [64], and efficiency [65].

In addition, one of the fundamental analyses of complex networks is community detection (also called clustering graph), which aims to decompose the network in order to find densely connected structures, so-called communities [66–68]. However, the detected communities have to be transformed into one scalar value in order to include them into the machine working workflow. Therefore, we defined new measures by first detecting all communities, then selecting the largest one and calculating its average path length. For the detection of community structures, different algorithms from the literature were used: Fastgreedy community (FC) [69], infomap community (IC) [70], leading eigenvector community (LC) [71], label propagation community (LPC) [72], edge betweenness community (EBC) [73], spinglass (SPC) [74] and multilevel community (MC) [75]. Since the community detection algorithms were combined with the average path length, we extended the abbreviations by the letter “A” as follows: AFC, AIC, ALC, ALPC, AEBC, ASPC and AMC.

Figure 3 depicts the entire workflow.

## IV. MACHINE LEARNING ALGORITHM

### A. Classification

In order to classify the (A) EEG time series, (B) the connectivity matrices, and (C) the complex network measures, the support vector machine (SVM) [76] algorithm was used. SVM has been used with excellent results for the classification of complex network measures before by other groups [77–79] and performed superior in our comparative evaluation. In this analysis, we compared the following machine learning methods to classify the complex network measures: Random forest (RF) [80], SVM [76], naive bayes (NB) [81], multilayer perceptron (MLP) [82], stochastic gradient descent with linear models classifier (SGD) [83], logistic regression (LG) [84] and extreme Gradient Boosting classifier [85] (XGBoost). The results can be found in Appendix C.

### B. Resampling and evaluation

The dataset was resampled by separating it into training (train) and test sets, with 25% of data composing the test set. Then, for a reliable model, a k-cross validation was used [86], with k = 10 (value widely used in the literature [87–91]). A hyper-parameter optimization called grid search was used here, similar to [92–96]. The hyper-parameter optimization values used for each classifier models can be found in Appendix C.

For evaluation, the standard performance metrics of the accuracy (Acc.) was used as state-of-art in literature [97–101]. Since the problem here is a two-class (negative and positive) classification problem, another metrics considered here are the measures of precision and recall also used in the literature [102–105]. Precision (also called specificity) corresponds to the hit rate in the negative class (here corresponding to control or no effect related to ayahuasca). Whereas recall (also called sensitivity) measures how well a classifier can predict positive examples (hit rate in the positive class), here related with an effect of the ayahuasca. Another measure used here and also used in literature [93, 106, 107] is the F1 score which is the harmonic mean of the recall and precision [108]. For visualization of these two latter measures, the receiver operating characteristic (ROC) curve is a common method as it displays the relation between the rate of true positives and false positives. The area below this curve, called area under ROC curve (AUC) has been widely used in classification problems [95, 97, 109, 110]. The value of the AUC varies from 0 to 1, where the value of one corresponds to a classification result free of errors. *AUC* = 0.5 indicates that the classifier is not able to distinguish the two classes, this result is equal to the random choice. Furthermore, we consider the micro average of ROC curve, which computes the AUC metric independently for each class (calculate AUC metric for healthy individuals, class zero, and separately calculate for unhealthy subjects, class one) and then the average is computed considering these classes equally. The macro average is also used in our evaluation, which does not consider both classes equally, but aggregates the contributions of the classes separately and then calculates the average.

Furthermore, we interpret the machine learning results using SHapley Additive exPlanations (SHAP) values [111] to quantify the importance of the complex measures, connections of brain regions, and location of electrodes for the classification result. This metric enables the identification and prioritization of features and can be used with any machine learning algorithm [112–114].

## V. RESULTS

The highest classification performance was obtained using the connectivity matrices with an accuracy of 92%, followed by the EEG time series (88%) and the complex network measures (83%) (see Table II). The following subsections V A, V B and V C contain the results in more detail.

**TABLE II:** Performances of the SVM classifier on the different types of data used in this paper. The best performance is highlighted in bold. The classification of connectivity matrices best captured the changes in the brain due to ayahuasca.

| Type of data | Subset | AUC | Acc. | F1 score | Recall | Precision |
| --- | --- | --- | --- | --- | --- | --- |
| EEG time series | Train | 0.87 | 0.89 | 0.88 | 0.87 | 0.89 |
|  | Test | 0.85 | 0.88 | 0.86 | 0.85 | 0.86 |
| Connectivity matrix | Train | **0.92** | **0.94** | **0.93** | **0.92** | **0.96** |
|  | Test | **0.88** | **0.92** | **0.90** | **0.88** | **0.94** |
| Complex measure | Train | 0.79 | 0.81 | 0.79 | 0.79 | 0.78 |
|  | Test | 0.75 | 0.83 | 0.78 | 0.75 | 0.90 |

### A. EEG time series

The performance of the test sample using the EEG time series was mean AUC of 0.85, mean precision of 0.88, mean F1 score of 0.86, mean recall of 0.85, and mean accuracy of 0.86. The precision measure is related to the positive class (with ayahuasca). Since the precision measure was slightly higher than the recall measure, the model can better detect the presence of ayahuasca instead of the absence of it.

In Figure 4, the confusion matrix (4-(a)), the learning curve (Figure 4-(b)), and the ROC curve (4-(c)) are plotted. The learning curve evaluates the predictability of the model by varying the size of the training set [114]. Figure 4-(b) shows that the highest accuracy in the test sample can only be achieved when the entire database is used.

**FIG. 4:**
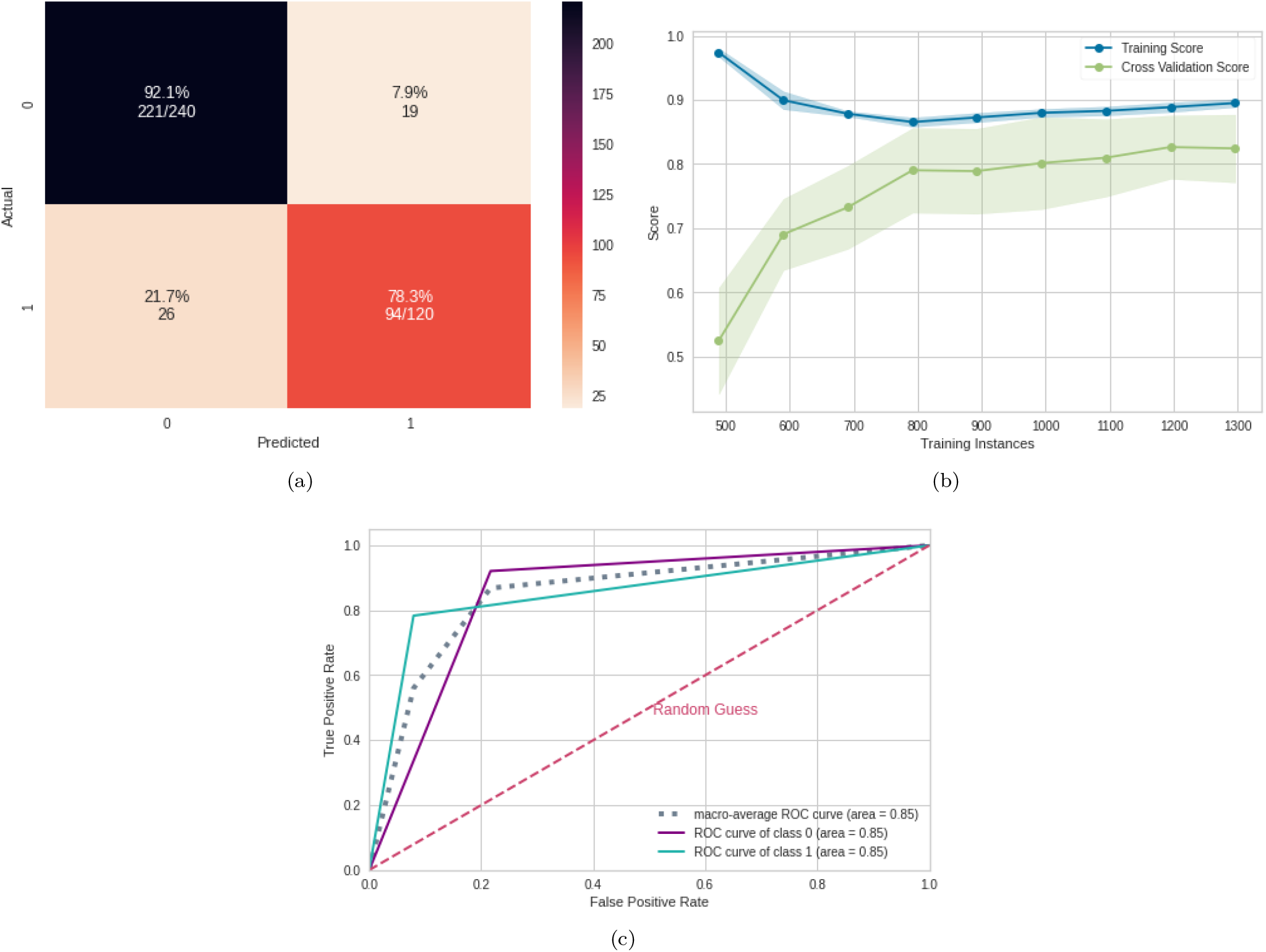
Machine learning results using the EEG time series as input data. (a) Confusion matrix indicating a true negative rate of 92.1% (blue according to the color bar) and a true positive rate of 78.3% (orange according to the color bar). (b) Learning curve for the training accuracy (blue) and for test accuracy (green). (c) ROC curve of class 0 (without ayahuasca) and class 1 (with ayahuasca). The gray dotted curve is the macro-average accuracy (area under curve = 0.85) and the pink one the random classifier.

Not all electrodes of the EEG recording were equally important for the classification. According to the SHAP values, the most important region for the model was T7, located in the temporal region (see Figure 5). This region was followed, in order of importance, by FC1, Fp1, P5 and Fz, located in the region between frontal and central, between frontal and parietal, parietal and frontal, respectively (see Figure 6-(a)). In addition, Figure 6-(b) shows details of the impact of each feature on the model. Positive SHAP values are shown when the presence of ayahuasca was detected, and negative SHAP values are shown when the absence of ayahuasca was detected. The colors indicate whether the feature value was low (blue) or high (red). Since the feature consists of the amplitudes of the EEG time series, it can be seen that for T7, the low amplitudes (blue dots) were important to detect the absence of ayahuasca (negative SHAP values), and the high amplitudes (red dots) were important to detect the presence of ayahuasca (positive SHAP values).

**FIG. 5:**
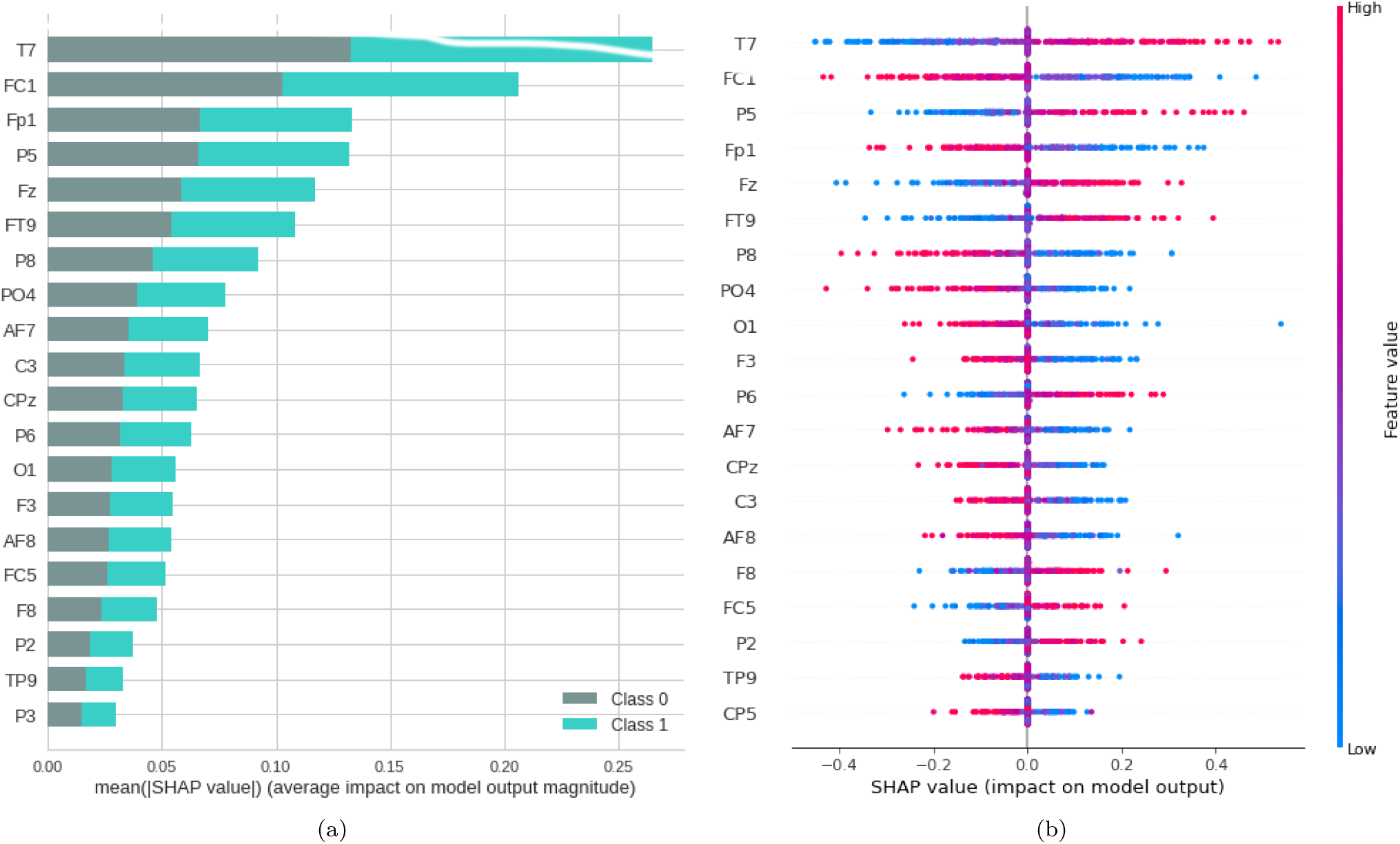
Feature importance ranking for SVM classifier being the brain regions ranked in descending order of importance. Brain region T7 is most important to classify the effect of ayahuasca. (a) Feature ranking based on the average of absolute SHAP values over all subjects considering both classes (gray:without ayahuasca, cyan: with ayahuasca). (b) Same as (a) but additionally showing details of the impact of each feature on the model.

**FIG. 6:**
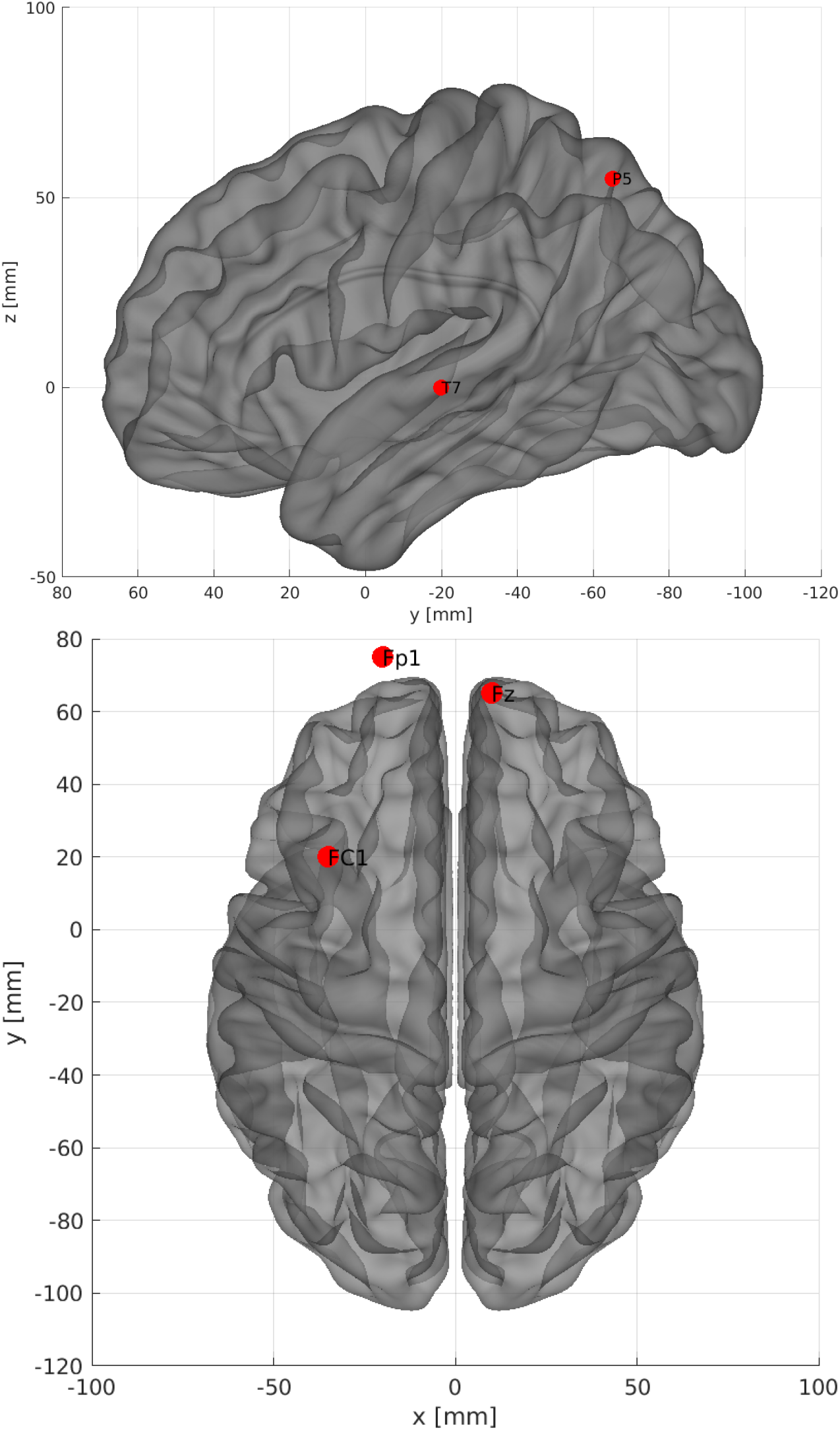
The five most important brain regions considering EEG time series as input data. (A) -Sagittal left plane showing the brain region for the channel T7 and P5. (B) Axial dorsal plane showing the brain regions Fz, Fp1 and FC1. The brain plot was made using Braph tool [115], based on the coordinates in [116, 117].

### B. Connectivity matrices

For the connectivity matrices, the test sample performance was a mean AUC of 0.88, mean accuracy of 0.92, mean F1 score of 0.90, mean recall of 0.88, and mean precision of 0.94.

Similar to the previous subsection V A, the precision measure was higher than the recall measure and therefore the model can better detect the presence of ayahuasca. In Figure 7, the confusion matrix (Figure 7-(a)), the learning curve (Figure 7-(b)), and the ROC curve (Figure 7-(c))are plotted. Similar to EEG time series, the learning curve for the connectivity matrices shows that the highest accuracy in the test sample can only be achieved when the entire database is used.

**FIG. 7:**
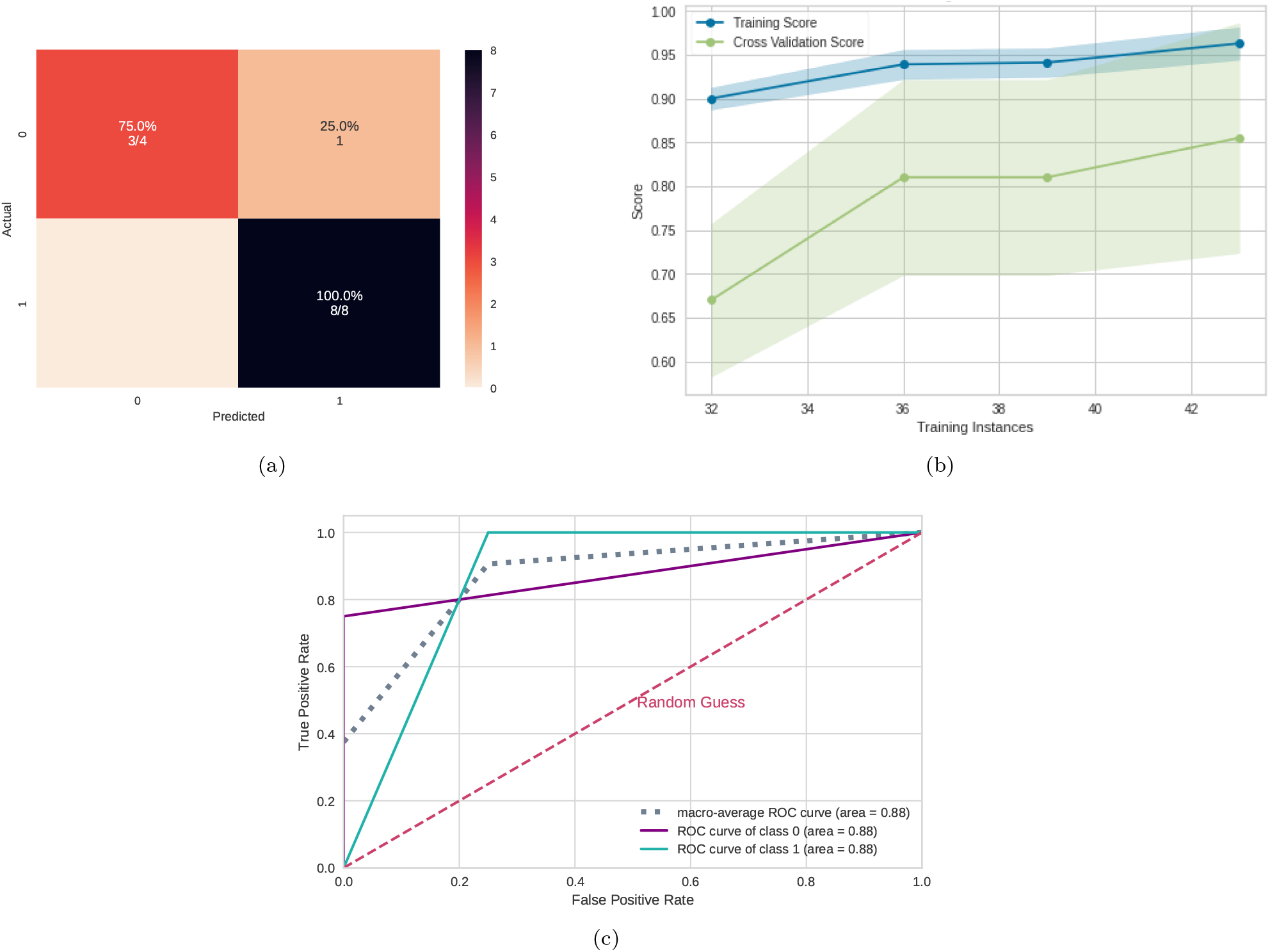
Machine learning results using the connectivity matrices as input data. (a) Confusion matrix indicating a true negative rate of 75% (orange according to the color bar) and a true positive of 100% (blue according to the color bar). (b) Learning curve for the training accuracy (blue) and for test accuracy (green). (c) ROC curve of class 0 (without ayahuasca) and class 1 (with ayahuasca). The gray dotted curve is the macro-average accuracy (area under curve = 0.88) and the pink one the random classifier.

In order to reveal the importance of the brain connections, the SHAP values were used as in the previews subsection. The results are shown in Figure 8. From that the most important connection was between F3 (frontal region) and PO4 (between parietal and occipital region). In addition, in Figure 8-(b) it can be seen that for the connection between F3 and PO4, low values of correlation (blue dots) were important for detecting the absence of ayahuasca (negative SHAP values), and high values of correlation (red dots) were important for detecting the presence of ayahuasca (positive SHAP values).

**FIG. 8:**
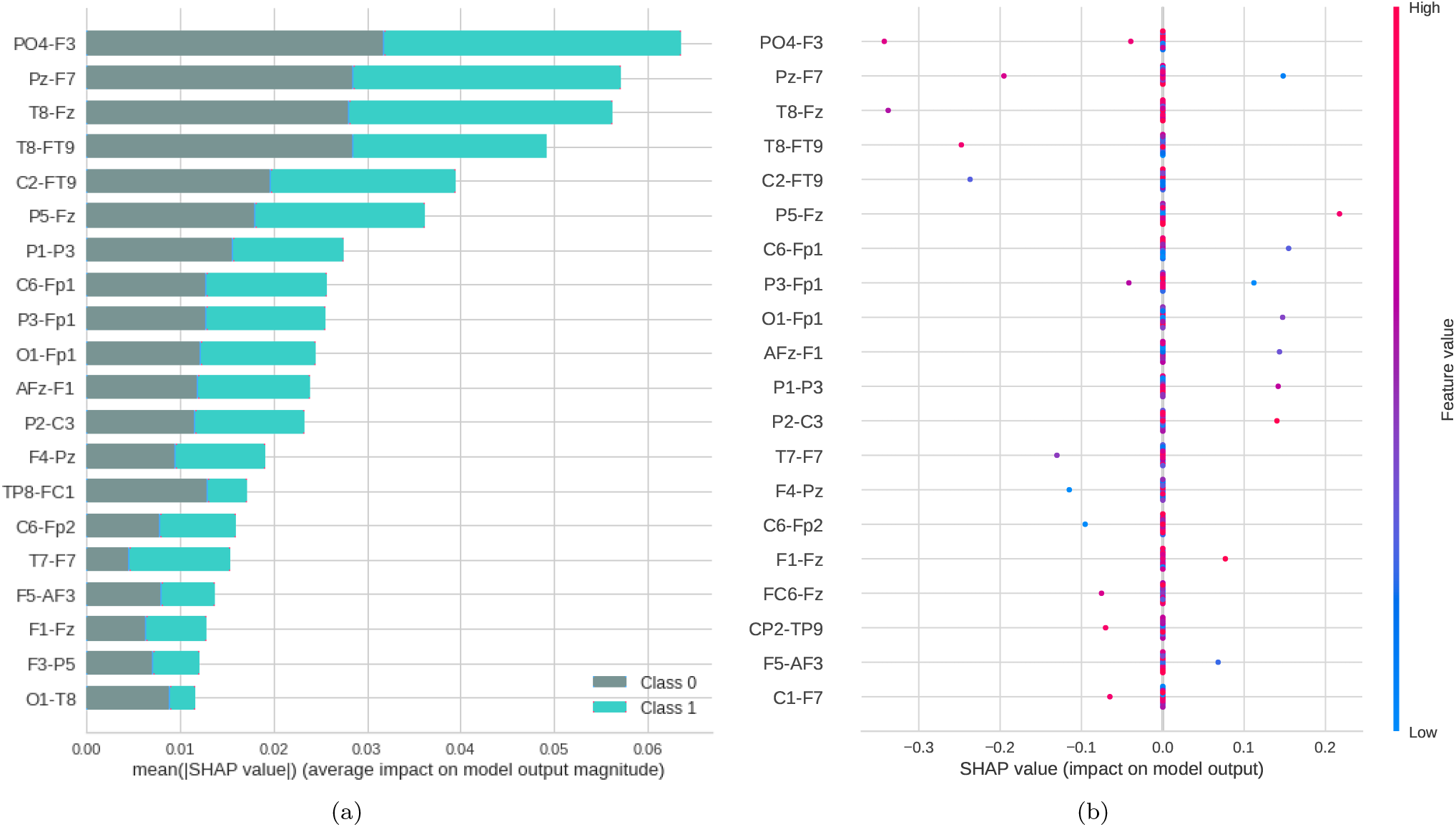
Feature importance ranking for SVM classifier being the connections of brain regions ranked in descending order of importance. The connection between the regions PO4 and F3 is the most important to classify the effect of ayahuasca. (a) Feature ranking based on the average of absolute SHAP values over all subjects considering both classes (gray: without ayahuasca, cyan: with ayahuasca). (b) Same as (a) but additionally showing details of the impact of each feature on the model.

The location in the brain can be seen on Figure 9.

**FIG. 9:**
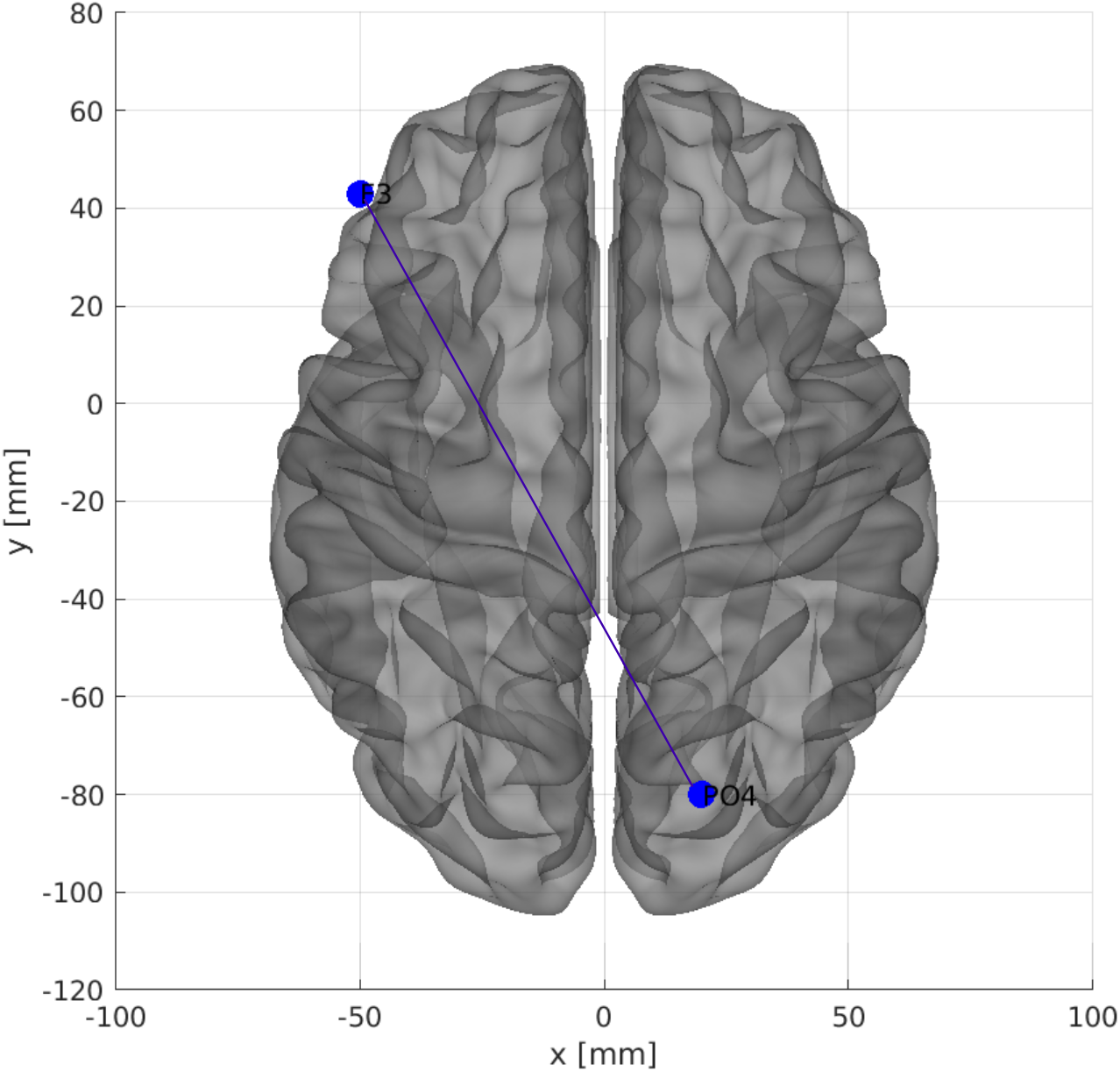
The most important connection of brain regions considering connectivity matrices as input data. Axial dorsal plane showing the brain regions connection between F3 and PO4. The brain plot was made using Braph tool [115], based on the coordinates in [116, 117].

### C. Complex network measures

The test sample performance using the complex network measures was a mean AUC of 0.75, mean accuracy of 0.83, mean F1 score of 0.78, mean recall of 0.75, and mean precision of 0.90.

Similar to the previous subsections V A and V B, the precision measure was higher than the recall measure and therefore the model can better detect the presence of ayahuasca.

In Figure 10, the confusion matrix (Figure 10-(a)), the learning curve (Figure 10-(b)), and the ROC curve (Figure 10-(c)) are plotted. Again, the entire database is necessary in order to get the highest accuracy.

**FIG. 10:**
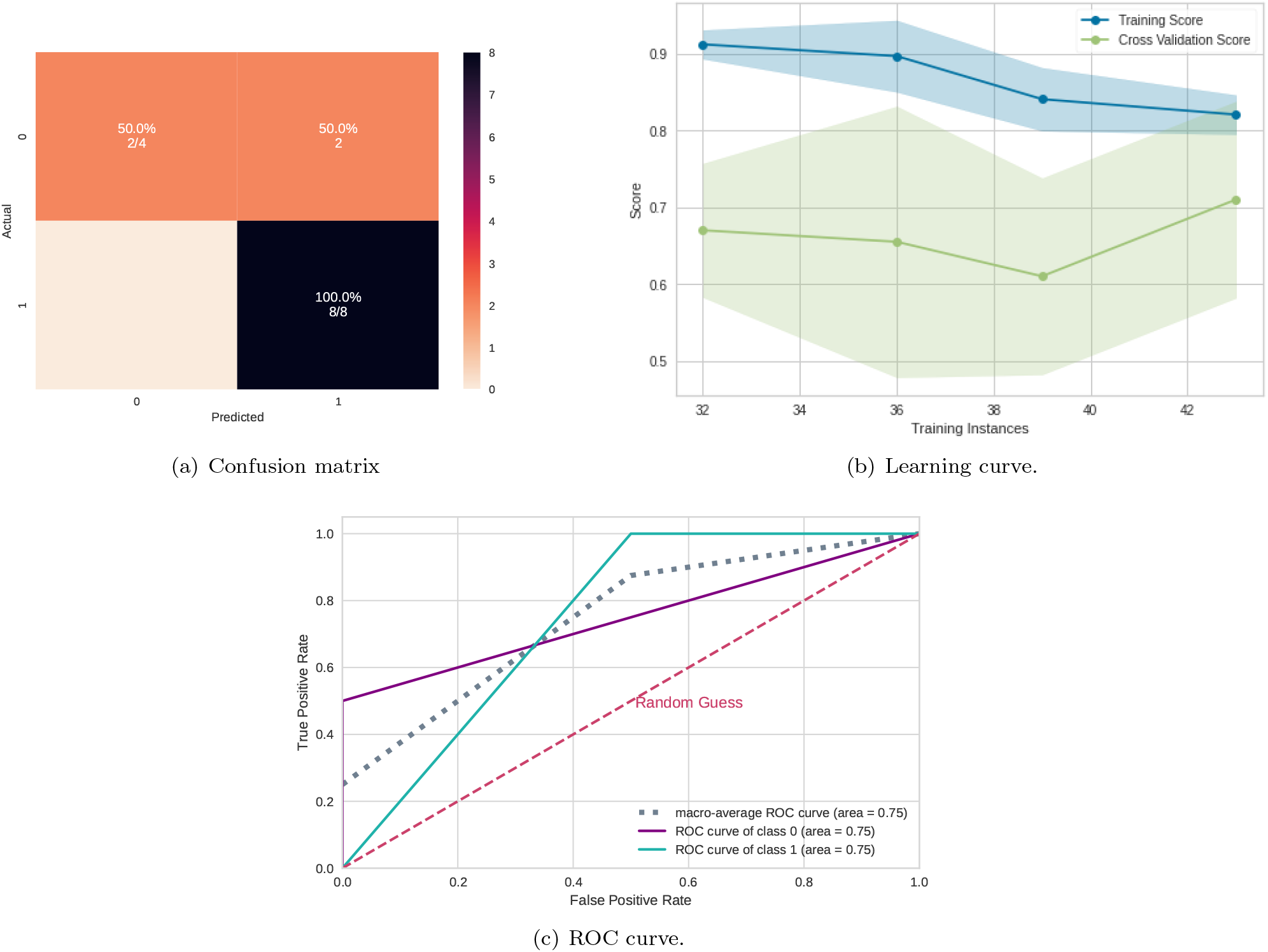
Machine learning results using the complex network measures as input data. (a) Confusion matrix indicating a true negative rate of 50% (orange according to the color bar) and a true positive rate of 100% (blue according to the color bar). (b) Learning curve for the training accuracy (blue) and for test accuracy (green). (c) ROC curve of class 0 (without ayahuasca) and class 1 (with ayahuasca). The gray dotted curve is the macro-average accuracy (area under curve = 0.75) and the pink one the random classifier.

From the SHAP values in Figure 11 it can be seen that the most important measure for the model was the CC, followed by assortativity, and the newly introduced measures ASC and ASPC. In addition, in Figure 11-(b) can be seen that for the CC measure, low values of this metric (blue dots) were important for detecting the absence of ayahuasca (negative SHAP values), and high values of this metric (red dots) were important for detecting the presence of ayahuasca (positive SHAP (c) values).

**FIG. 11:**
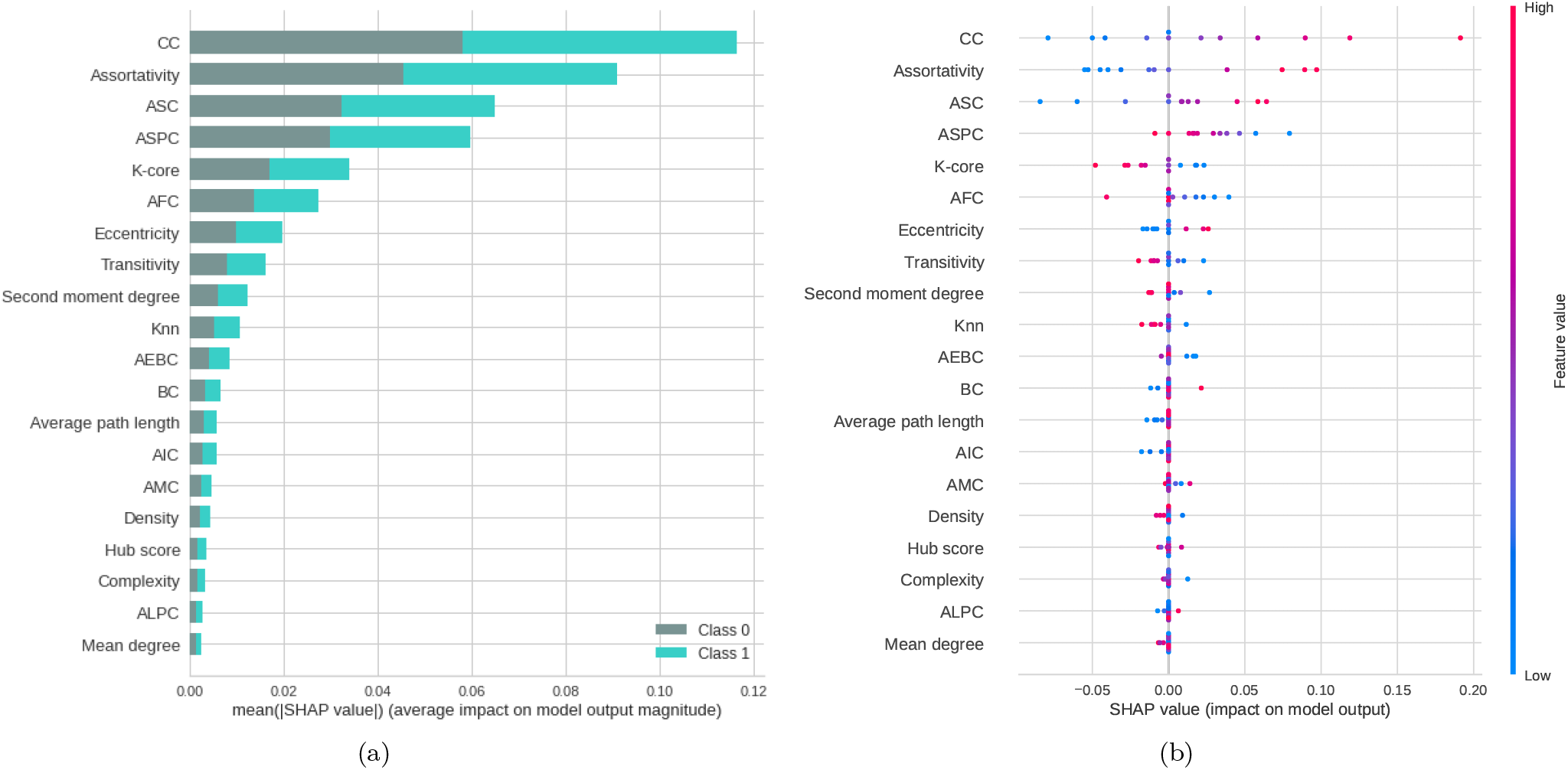
Feature importance ranking for SVM classifier being the features ranked in descending order of importance. The CC measure is the most important to classify the effect of ayahuasca. (a) Feature ranking based on the average of absolute SHAP values over all subjects considering both classes (gray: without ayahuasca, cyan: with ayahuasca). (b) Same as (a) but additionally showing details of the impact of each feature on the model.

## VI. DISCUSSION

In this paper, we aimed to answer the question if it is possible to automatically detect the brain activity changes due to ayahuasca using machine learning and which features are most important and could act as biomarkers.

Our results show that it is possible to automatically detect the changes due to ayahuasca. The classification accuracy was above 75% for all three data abstraction levels. The classification accuracy of connectivity matrices was higher than the raw EEG time series, suggesting that connection changes are more important between brain regions than within brain regions. This result is important, since the connectivity matrices not only improved the accuracy, but also produced efficiency gains, such as reduced data storage and faster machine learning training. This would be especially useful for larger datasets, where raw time series may be very costly, for example in hospital diagnosis systems.

### A. EEG time series

The analysis of the raw EEG time series revealed that the frontal and the temporal lobe were the most affected brain regions. In line with that, studies using single photon emission computed tomography (SPECT) have reported that ayahuasca increases blood perfusion in the frontal regions of the brain, more specifically the insula, left nucleus accumbens, left amygdala, parahippocampal gyrus, and left subgenual area [16, 118]. Furthermore, works using functional magnetic resonance imaging have observed activation in the occipital, temporal, and frontal areas of the brain [10, 119]. These regions are related to introspection, emotional processing and in the therapeutic effects of traditional antidepressants [120] and most interestingly, it may also affect motor and cognitive functions in other neurological disorders, such as Parkinson’s disease and Alzheimer’s disease, respectively [121, 122].

### B. Connectivity matrices

In terms of brain connections, the correlation between left frontal cortex (F3) and right parietal-occipital (PO4) was most important. [123] showed that synchronization in the gamma band between the parietal-occipital and frontal cortices was present during tasks of face recognition. Since the EEG time series data used in this work only contained the gamma band, the P04-F3 connection could point to a similar cognitive processes in the subjects during ayahuasca mediated visual hallucinations.

### C. Complex network measures

The most important complex network measure was CC. CC is a centrality measure which can be defined as the inverse of the average length of the shortest path from one node to all other nodes in the network [124]. The idea is that important nodes participate in many shortest paths within a network and, therefore, plays an important role in the flow of information in the brain [51]. The CC was also the most important measure in other papers related to the differentiation of patients with AD (c) [125–128]. In these papers, CC was shown to decrease due to AD disease, while ayahuasca ingestion increased the median value of this measure (see on the Figure 12-(a)).

**FIG. 12:**
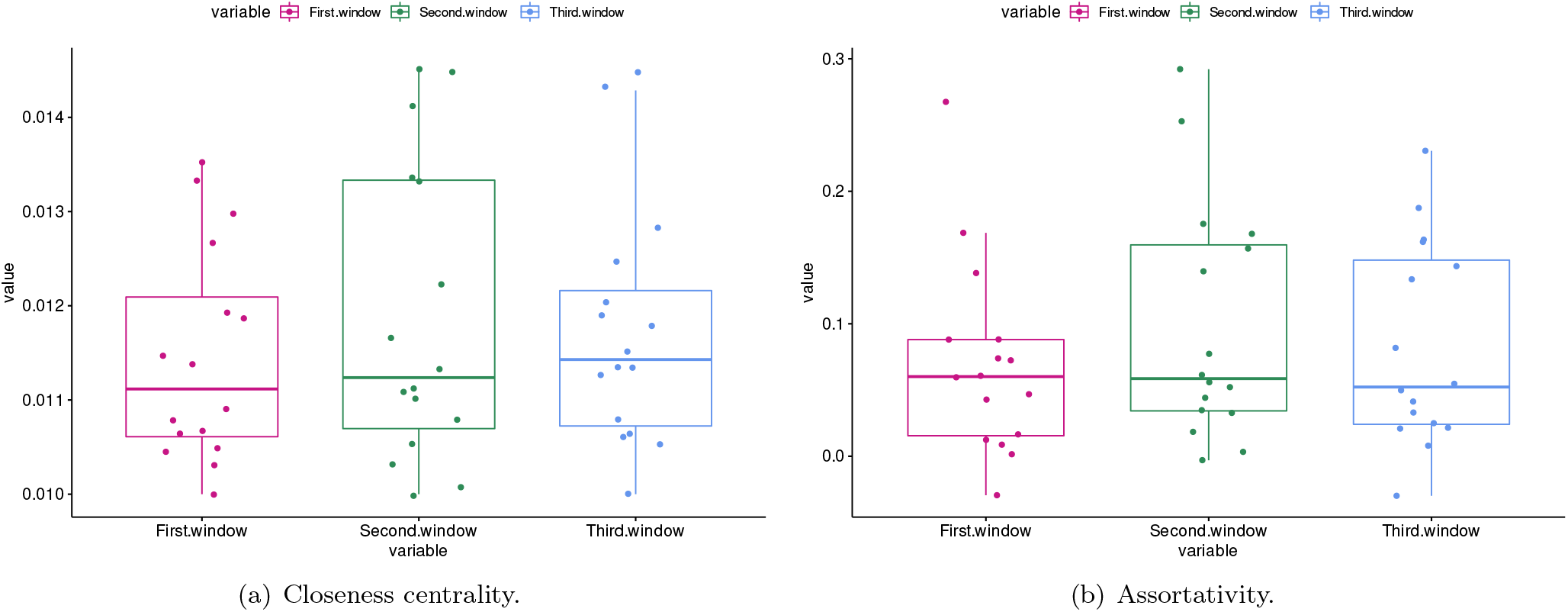
Boxplot of the closeness centrality and assortativity measures for all subjects in the first, second and third windows (respectively in pink, green and blue). It can be seen that the median of the closeness centrality measure (central bar in the boxplot) increased with the use of ayahuasca. The median of the assortativity, in contrast, decreased with the use of ayahuasca.

The second most important complex network measure was the assortativity. This measure refers to the resilience of networks [48]. A positive assortativity coefficient indicates a network with a resilient core due to the interconnected nodes of high degree [124]. This measure was also associated with AD in several works [129, 130] whose results showed an increase in the assortativity value in contrast to what was found here, where with the use of ayahuasca the assortativity value (median) decreased (see on the Figure 12-(b)). It should be noted that although the median value decreased, the upper confidence interval of the distribution increased.

In summary, the results suggest a possible relationship between ayahuasca and AD in terms of brain network, indicating a therapeutic potential. Indeed, a possible mechanism of how ayahuasca acts against AD was described in [19]. According to this, the ayahuasca compound dimethyltryptamine (DMT) agonizes the sigma 1 receptor (Sig-1R) and thereby regulates endoplasmic reticulum (ER) stress and Unfolded Protein Response (UPR), which are thought to play a key role in neuropsychiatric diseases such as AD.

The third most important complex network measure was ASC. It quantifies the average path length of the largest community. In this context, the average path length measures the efficiency of information transfer within the community. To the best of our knowledge, a measure such as ASC has not been used before.

Overall, the classification was successful by taking into account the complete set of measures, rather than just one single measure. Like shown in Figure 12, even the most important measures CC and assortativity did not show much difference between the first window (without ayahuasca) and the other windows (with ayahuasca). Together with the other less important measures, however, the machine learning method was able to successfully distinguish both classes. This leads to the conclusion that a single feature is insufficient as a biomarker, while the set of different features used in this work may serve as a biomarker.

## VII. CONCLUSIONS AND FUTURE WORK

In summary, the results obtained in our study demonstrated that the application of machine learning methods was able to automatically detect changes in brain connectivity during ayahuasca use.

The findings also suggest that the use of this substance affects important brain regions related to different cognitive, psychiatric, and motor functions. These effects may alleviate different symptoms of diseases affecting the brain. Future studies are needed to better understand its neurochemical and long-term effects on patients’ health. Finally, the same methodology applied here may be useful in interpreting EEG time series from patients who consumed other psychedelic drugs, such as pure DMT [131]. In future work, we aim to apply this workflow to recordings from our laboratory using in vitro neuronal networks on microelectrode arrays to study the effects of psychedelics at a single network level.

Thus, regardless of the equipment used to collect the data, we would like to verify whether the same method used here is able to detect changes due to different psychedelics.

## Data Availability

All data produced are available online at https://dataverse.harvard.edu/dataset.xhtml?persistentId=doi:10.7910/DVN/VVE6QC

## VIII. ACKNOWLEDGEMENTS

F.A.R. acknowledges CNPq (grant 309266/2019-0) and FAPESP (grant 19/23293-0) for the financial support given for this research. A.M.P. acknowlwdges FAPESP (grant 2019/22277-0) for the financial support given this research. KR acknowledges FAPESP grant 2019/26595-7.

## Appendix A: EEG database electrodes

**FIG. 13:**
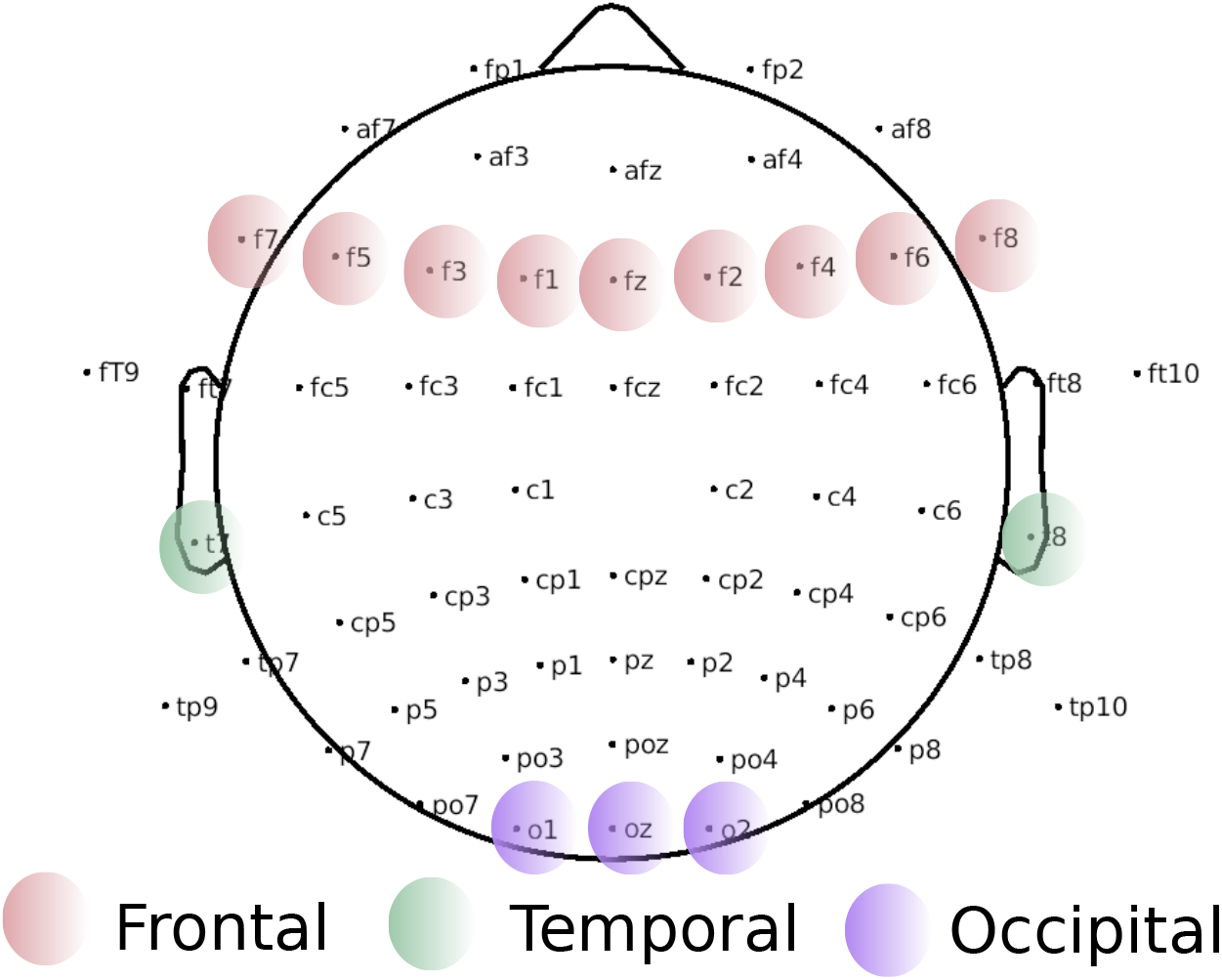
Head model for all electrodes used in the present work. The brain regions modulated after using ayahuasca, according to literature (see section I), are the frontal, temporal, and occipital lobes, highlighted in the Figure in pink, green and purple, respectively. Developed by the authors using MNE-Python [132]

## Appendix B: Hyperparameter values of Grid Search classifier

**TABLE III:**
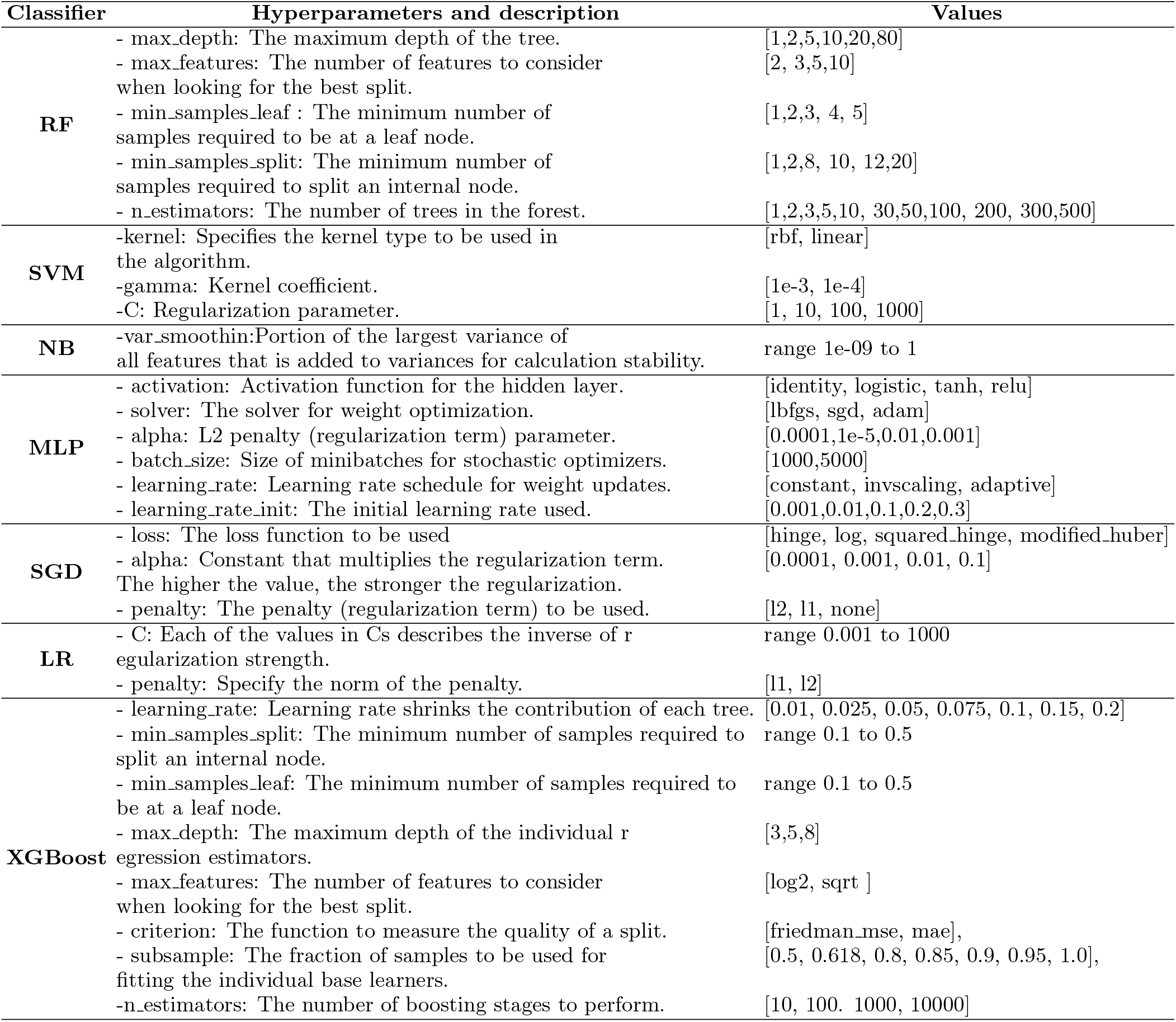
Table containing the hyperparameters for each classifier using the Grid search optimizer.

## Appendix C: Results of all classifier in the complex network measures

**TABLE IV:**
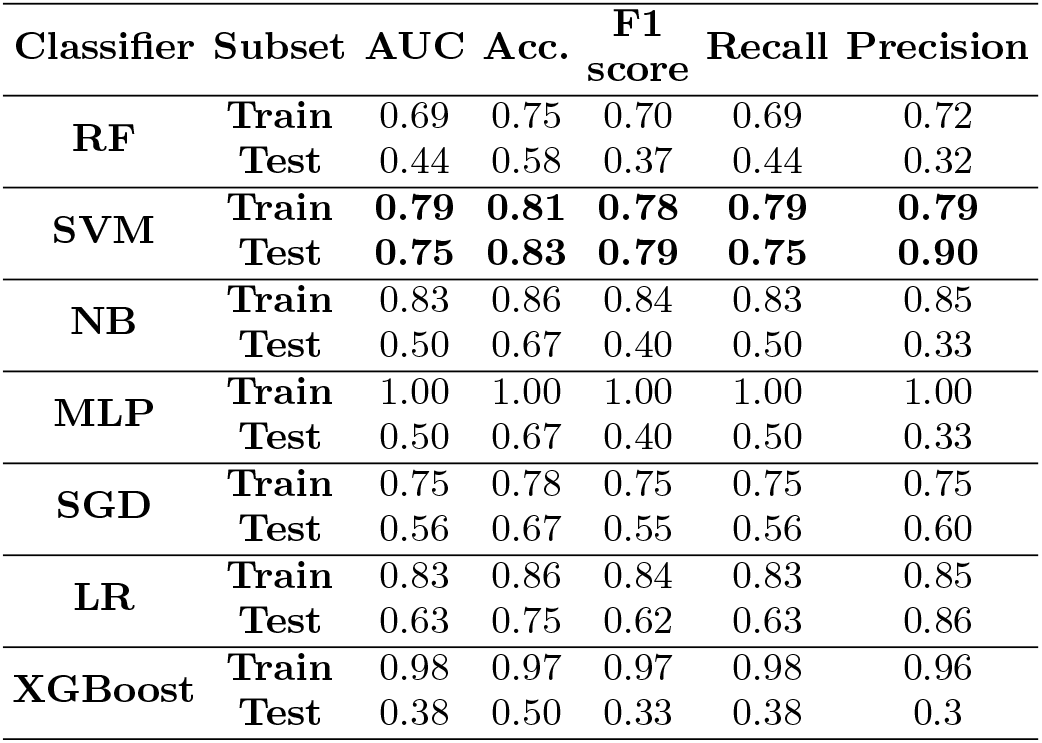
Performance of all classifiers applied to the network measurements. In bold the best performance referring to the SVM classifier.

| Classifier | Subset | AUC | Acc. | F1<br>score | Recall | Precision |
| --- | --- | --- | --- | --- | --- | --- |
| RF | Train | 0.69 | 0.75 | 0.70 | 0.69 | 0.72 |
|  | Test | 0.44 | 0.58 | 0.37 | 0.44 | 0.32 |
| SVM | Train | **0.79** | **0.81** | **0.78** | **0.79** | **0.79** |
|  | Test | **0.75** | **0.83** | **0.79** | **0.75** | **0.90** |
| NB | Train | 0.83 | 0.86 | 0.84 | 0.83 | 0.85 |
|  | Test | 0.50 | 0.67 | 0.40 | 0.50 | 0.33 |
| MLP | Train | 1.00 | 1.00 | 1.00 | 1.00 | 1.00 |
|  | Test | 0.50 | 0.67 | 0.40 | 0.50 | 0.33 |
| SGD | Train | 0.75 | 0.78 | 0.75 | 0.75 | 0.75 |
|  | Test | 0.56 | 0.67 | 0.55 | 0.56 | 0.60 |
| LR | Train | 0.83 | 0.86 | 0.84 | 0.83 | 0.85 |
|  | Test | 0.63 | 0.75 | 0.62 | 0.63 | 0.86 |
| XGBoost | Train | 0.98 | 0.97 | 0.97 | 0.98 | 0.96 |
|  | Test | 0.38 | 0.50 | 0.33 | 0.38 | 0.3 |

## Footnotes

1 Avaiable on [38].

